# Persistent morbidity and knowledge gaps in a near-elimination setting: A cross-sectional study of lymphatic filariasis in northern Ghana

**DOI:** 10.64898/2026.04.21.26351358

**Authors:** Cosmas Ngmaanoma Ngmembong, Millicent Captain-Esoah, Sharmila Lareef, Elijah Dakorah Angyiereyiri, Martin Ntiamoah Donkor, Sylvester Donne Dassah, Mawuli Kwamla Azameti, Grace Adzo Motey, Lawrence Adelani Adetunde, Theophilus Atio Abalori, Kwadwo K. Frempong, Chrysantus Kubio, Daniel Adjei Boakye, Samuel Kweku Dadzie, Felicia Owusu-Antwi, Michael Rockson Adjei, Martin Peter Grobusch, Didier Bakajika, Frank John Lule, Fiona Braka, Dorothy Achu

## Abstract

**Background:** Lymphatic filariasis remains a public health concern in many endemic regions, where chronic disease persists despite substantial reductions in transmission. In Ghana, more than two decades of mass drug administration have significantly reduced disease prevalence and transmission; however, chronic manifestations and gaps in community understanding continue to be reported in parts of the north. This study assessed infection status, chronic morbidity burden, and community knowledge in a rural setting in northern Ghana approaching elimination.

**Methodology/Principal Findings:** A community-based cross-sectional study was conducted in Birifor, northern Ghana, from October 2024 to January 2025. A total of 261 residents aged ten years and above were selected using random sampling. Data collection included structured questionnaires, clinical examination for chronic disease, and night blood microscopy for the detection of infection.

No microfilariae were detected (0/261; 0%). However, chronic lymphoedema was identified in five individuals (1.9%), all aged over 40 years. Awareness of the disease was high (95.8%), yet only 39.5% of participants demonstrated good community knowledge and perceptions and self-reported preventive practices. Misconceptions regarding transmission, particularly beliefs that the disease is hereditary or caused by spiritual factors, were common. Participation in mass drug administration was high (93.1%). Despite this, chronic disease imposed a notable socioeconomic burden: all affected individuals reported loss of income, and 60% reported additional household income loss due to caregiving.

**Conclusions/Significance:** These findings suggest that transmission in the study area is likely very low, although residual infection cannot be excluded; however, chronic disease and gaps in community knowledge persist. Strengthening morbidity management, improving community education, and providing support for affected households are essential. Sustained surveillance and integrated approaches will be critical to prevent resurgence and support long-term elimination efforts.

**Author Summary:** Lymphatic filariasis, also known as elephantiasis, is a mosquito-borne disease that can cause long-term swelling of the legs, arms, or genitals. Global efforts have greatly reduced its occurrence, especially through repeated mass drug administration to afflicted communities. However, many people continue to live with chronic swelling caused by past infections, which can affect their ability to work and participate fully in daily life.

In this study, we examined the current situation of lymphatic filariasis in a rural community in northern Ghana that has received many years of treatment. We tested people for active infection, assessed signs of chronic disease, and explored what community members know and believe about the disease.

We found no evidence of active infection, suggesting that transmission is now very low. However, some individuals were still living with chronic swelling and reported loss of income, while households also experienced financial strain due to caregiving. Although most people had heard of the disease, many did not fully understand how it is transmitted.

Our findings show that reducing transmission alone is not enough. Continued education, community support, and access to care are needed to address the long-term impact of the disease and support ongoing elimination efforts.

## Background

Lymphatic filariasis (LF) is a mosquito-borne neglected tropical disease that can cause progressive lymphatic dysfunction, initially presenting with transient swelling and, if untreated, progressing to chronic lymphoedema, hydrocele, pain, disability, and long-term psychosocial and economic consequences for affected individuals and households [1–3]. Global elimination efforts accelerated after the launch of the Global Program to Eliminate Lymphatic Filariasis (GPELF) in 2000, which is built on two interdependent pillars: (i) annual mass drug administration (MDA) to reduce microfilariae levels and interrupt transmission, and (ii) morbidity management and disability prevention (MMDP) to improve quality of life for people living with chronic disease [4–6]. Many endemic countries have since achieved substantial declines in microfilaria prevalence and vector infection rates, and several have been validated as having eliminated LF as a public health problem [5].

Despite these gains, programmatic challenges persist in areas with ongoing transmission or residual chronic morbidity. Evidence from multiple settings shows that even after microfilaria prevalence approaches zero, individuals may continue to present with lymphoedema or hydrocele due to past infections, gaps in access to care, or limited awareness of preventive practices [6–9]. Persistent pockets of transmission have been documented in several countries, where district-level heterogeneity in morbidity trends remains despite overall declines [10,11]. These insights underscore the importance of integrating epidemiologic surveillance with social and behavioural research to guide targeted, context specific interventions.

Ghana has implemented LF MDA for more than two decades and has stopped community-wide treatment in most previously endemic districts [8,12]. However, persistent antigen positivity and reports of chronic disease in parts of northern Ghana suggest the need for continued surveillance and community-based assessments [13]. Studies from northern Ghana have reported higher prevalence of microfilaraemia, hydrocele, lymphoedema, and skin manifestations compared to southern zones, pointing to geographic heterogeneity in both infection and morbidity patterns [14]. For example, Adu-Mensah et al. documented Og4C3 antigen prevalence exceeding the WHO threshold for stopping MDA in some districts of the Upper East Region, suggesting ongoing transmission despite years of intervention [8,15]. Understanding how communities perceive causes, symptoms, transmission, prevention behaviours, and MDA campaigns is therefore essential for sustaining programmatic progress and preventing recrudescence.

At the same time, confirmatory diagnostics remain an area of active debate in elimination settings. While antigen detection assays and molecular xenomonitoring increase sensitivity for low level infections, operational costs and logistical barriers often limit their routine use in district health systems [16,17]. Similarly, long-term morbidity management remains under prioritized in many endemic regions despite evidence that simple limb care practices can reduce acute dermatolymphangioadenitis episodes and slow progression of lymphoedema [6,18,19]. These gaps highlight the need for integrated approaches that address both elimination and patient centred care.

Against this background, we conducted a community-based study in Birifor, a rural settlement along the Black Volta River in northern Ghana to (i) determine current infection status using night blood microscopy, (ii) describe the prevalence and severity of chronic disease through clinical examination, and (iii) evaluate community knowledge and perceptions and self-reported preventive practices regarding LF transmission, prevention, treatment, and care. We also explored how knowledge varied by demographic and socioeconomic characteristics. This study provides insights relevant for districts approaching elimination thresholds and highlights persistent knowledge gaps that may affect uptake of preventive measures and MDA participation.

## Methods

### Ethical Considerations

Ethical approval was obtained from the Navrongo Health Research Centre Institutional Review Board (NHRCIRB; reference number: NHRCIRB658, dated 8 October 2024). Additional permissions were obtained from the Lawra Municipal Health Directorate and community leadership.

Written informed consent was obtained from all adult participants. For participants under 18 years of age, assent was obtained in addition to consent from a parent or guardian. Participation was voluntary, and confidentiality of all participant information was strictly maintained.

### Study Site

Birifor is a rural community located in the Lawra Municipality of the Upper West Region of Ghana (10°30’02.3"N, 002°51’43.8"W). The community has an estimated population of approximately 3,500 residents. It lies about 2 km east of Babile and approximately 2 km west of the Black Volta River, which forms part of the international boundary between Ghana and Burkina Faso. See Fig 1. Most inhabitants are subsistence farmers engaged in mixed crop cultivation and livestock rearing.

**Fig 1:**
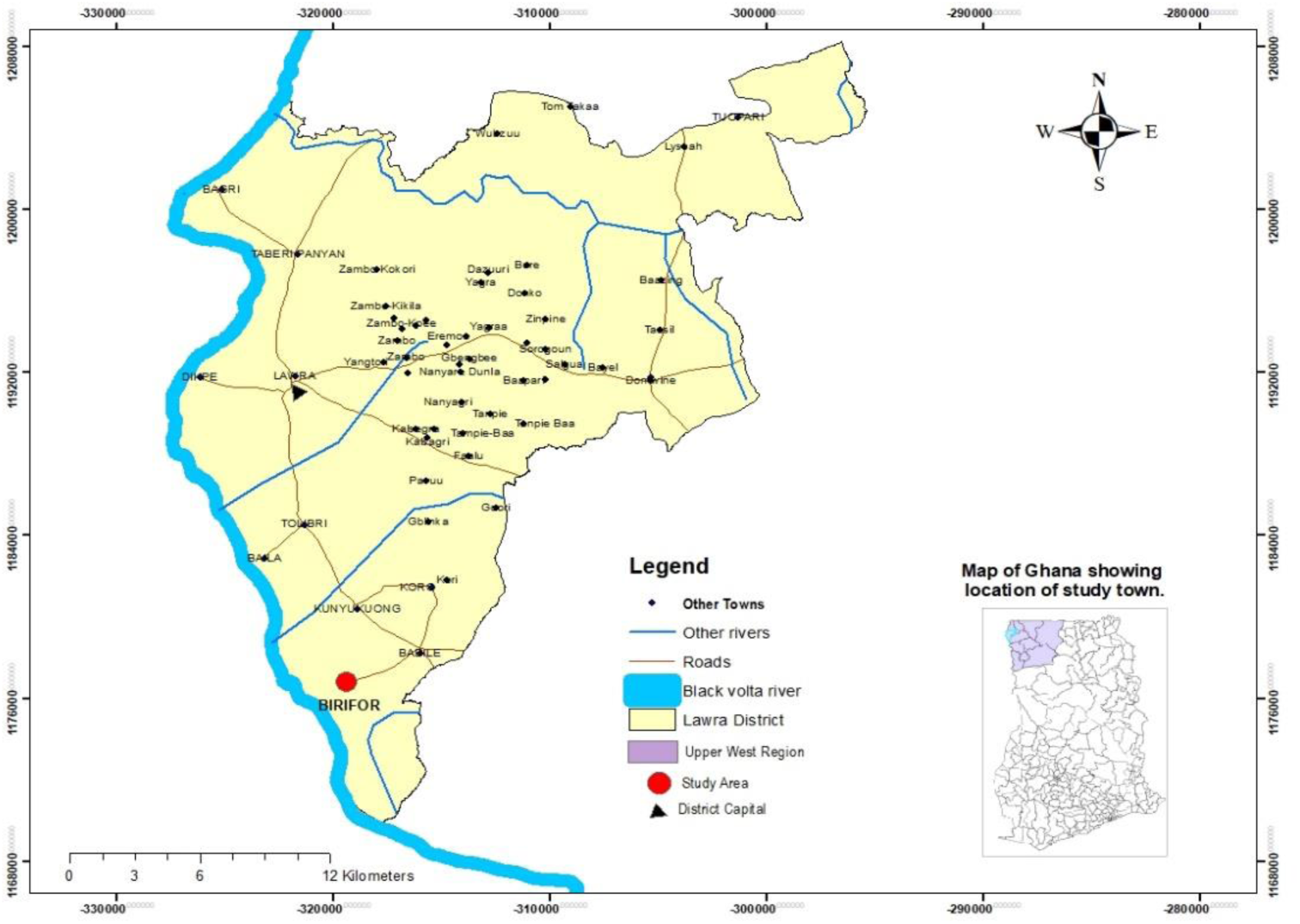
Map of Lawra Municipality.

### Study Design and Period

A community-based cross-sectional survey was conducted from 15 October 2024 to 20 January 2025 to assess (i) active *Wuchereria bancrofti* infection, (ii) chronic lymphatic morbidity, and (iii) community knowledge, attitudes, and practices (KAP) related to lymphatic filariasis (LF). All field procedures were implemented using standardized protocols consistent with WHO recommendations and approaches commonly used in LF elimination and morbidity studies published in PLOS NTD.

### Sample Size and Sampling Strategy

The sample size was determined using Cochran’s formula, assuming an expected prevalence of 50%, a 95% confidence level (Z = 1.96), and a margin of error of 5%. This yielded an initial sample size of 384 participants. After applying a finite population correction for the estimated population of 3,500, the minimum required sample size was 346. Due to logistical and operational constraints, 261 participants were enrolled. Although this was lower than the calculated minimum, the sample size was sufficient to detect key outcomes, including the absence of microfilaraemia and the presence of chronic morbidity, and provides useful insights into epidemiological and community-level patterns in this setting.

The most recent updated community census served as the sampling frame. Eligible individuals were assigned unique identification numbers, and simple random sampling was conducted using a computer-generated random number generator to select participants. In cases of non-response or ineligibility, replacement was performed using the same randomization procedure.

### Eligibility criteria

Individuals aged 10 years and above who had resided in Birifor for at least five years were eligible for inclusion.

Participants were excluded if they had taken antifilarial medications (diethylcarbamazine, ivermectin, or albendazole) within the preceding three months, declined to provide informed consent (or assent for minors), or were unwilling to provide night blood samples, undergo clinical examination, or complete the study questionnaire.

### Data Collection Procedures

Data were collected between 15 October 2024 and 20 January 2025 by trained field workers. Structured questionnaires were administered to collect socio-demographic information and selected indicators of community knowledge and self-reported preventive behaviors related to LF.

Clinical examinations were conducted by trained clinicians to identify signs of chronic LF manifestations, including lymphoedema and hydrocele.

Although the questionnaire included items on community knowledge and perceptions and self-reported preventive practices, the survey was not designed as a full knowledge attitude and practice (KAP) study, and the indicators presented here reflect a focused assessment of selected community knowledge and self-reported preventive behaviors rather than a comprehensive domain-specific KAP framework

### Parasitological assessment

Blood samples were collected from participants by certified medical laboratory scientists. Sampling was conducted at night between 22:00 and 02:00 hours to account for the nocturnal periodicity of microfilariae.

Both capillary and venous blood samples were obtained. Thin blood smears were prepared from capillary blood, while thick smears were prepared from venous blood samples. All smears were stained using standard procedures and examined microscopically for the presence of microfilariae. Only night blood microscopy was conducted; antigen detection assays or molecular methods were not available during fieldwork. Therefore, seroprevalence could not be assessed.

### Data management and analysis

Data collected using Kobo Collect were exported into Microsoft Excel and subsequently analyzed using STATA version 17 (StataCorp, College Station, TX, USA).

Descriptive statistics were used to summarize socio-demographic characteristics and qualitative responses. The indicators of knowledge and preventive behaviors were summarized using a simple classification adapted from previous LF studies. This was not intended as a full KAP evaluation but rather to provide contextual insight into community understanding and self-reported preventive actions Logistic regression analysis was performed to assess associations between KAP and socio-demographic variables. Crude odds ratios (cOR) with 95% confidence intervals (CI) were reported at the bivariate level, followed by multivariate analysis to adjust for potential confounders. Statistical significance was set at p < 0.05.

## Results

### Sociodemographic Characteristics of Participants

A total of 261 participants were enrolled in the study. Of these, 140 (53.7%) were male and 121 (46.3%) were female. Most participants were aged 10–30 years (161/261; 61.7%), with a mean age of 33.5 ± 16.7 years. Nearly half were unmarried (124/261; 47.5%), and 179 (68.6%) had attained primary education or less. Only seven participants (2.7%) reported being unemployed at the time of the survey. See Table 1.

**Table 1.**
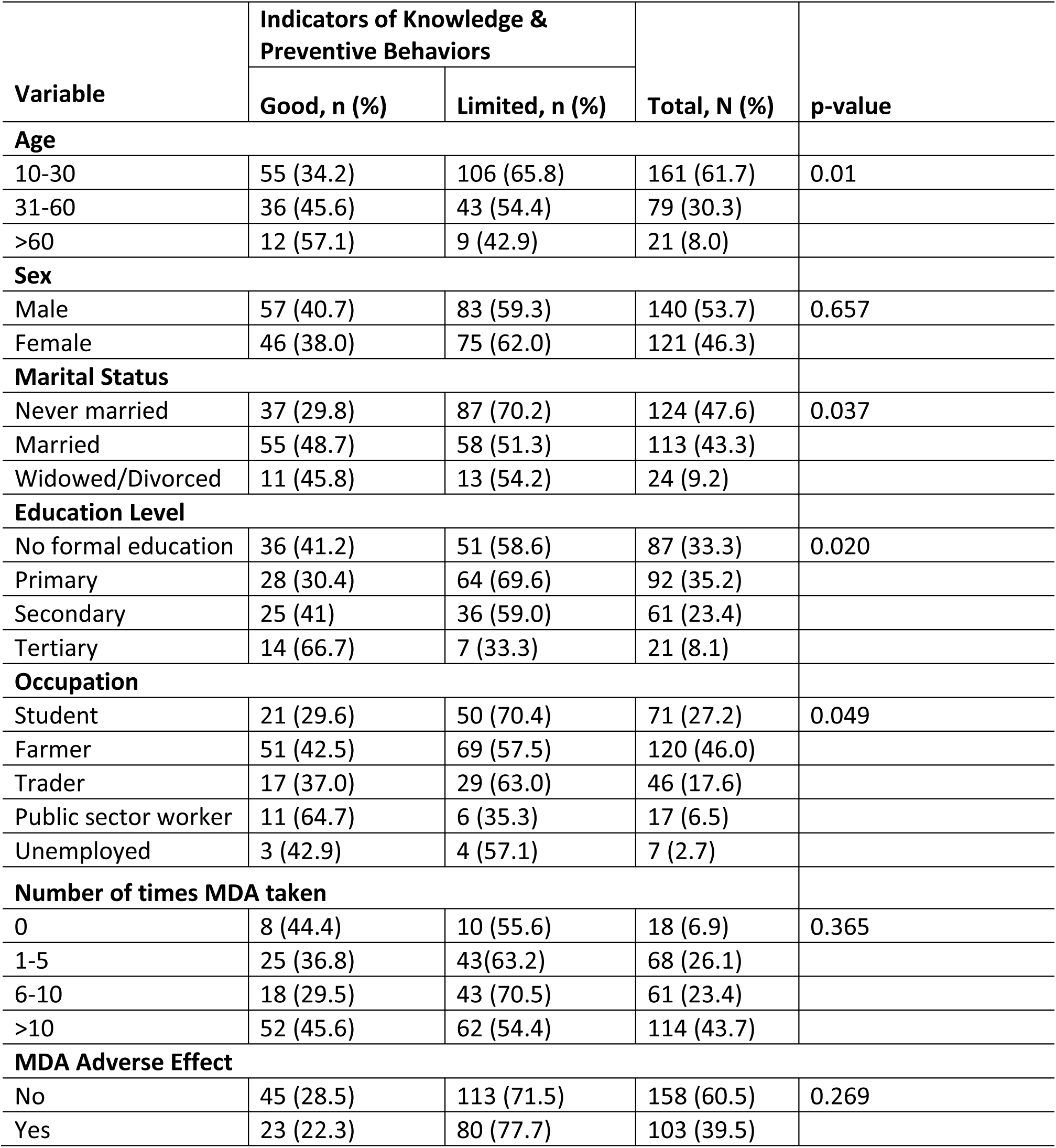
Sociodemographic characteristics and indicators of knowledge and preventive behaviors related to lymphatic filariasis among study participants in Birifor, Ghana.

| Variable | Indicators of Knowledge & Preventive Behaviors |  | Total, N (%) | p-value |
| --- | --- | --- | --- | --- |
|  | Good, n (%) | Limited, n (%) |  |  |
| **Age** |  |  |  |  |
| 10-30 | 55 (34.2) | 106 (65.8) | 161 (61.7) | 0.01 |
| 31-60 | 36 (45.6) | 43 (54.4) | 79 (30.3) |  |
| >60 | 12 (57.1) | 9 (42.9) | 21 (8.0) |  |
| **Sex** |  |  |  |  |
| Male | 57 (40.7) | 83 (59.3) | 140 (53.7) | 0.657 |
| Female | 46 (38.0) | 75 (62.0) | 121 (46.3) |  |
| **Marital Status** |  |  |  |  |
| Never married | 37 (29.8) | 87 (70.2) | 124 (47.6) | 0.037 |
| Married | 55 (48.7) | 58 (51.3) | 113 (43.3) |  |
| Widowed/Divorced | 11 (45.8) | 13 (54.2) | 24 (9.2) |  |
| **Education Level** |  |  |  |  |
| No formal education | 36 (41.2) | 51 (58.6) | 87 (33.3) | 0.020 |
| Primary | 28 (30.4) | 64 (69.6) | 92 (35.2) |  |
| Secondary | 25 (41) | 36 (59.0) | 61 (23.4) |  |
| Tertiary | 14 (66.7) | 7 (33.3) | 21 (8.1) |  |
| **Occupation** |  |  |  |  |
| Student | 21 (29.6) | 50 (70.4) | 71 (27.2) | 0.049 |
| Farmer | 51 (42.5) | 69 (57.5) | 120 (46.0) |  |
| Trader | 17 (37.0) | 29 (63.0) | 46 (17.6) |  |
| Public sector worker | 11 (64.7) | 6 (35.3) | 17 (6.5) |  |
| Unemployed | 3 (42.9) | 4 (57.1) | 7 (2.7) |  |
| **Number of times MDA taken** |  |  |  |  |
| 0 | 8 (44.4) | 10 (55.6) | 18 (6.9) | 0.365 |
| 1-5 | 25 (36.8) | 43(63.2) | 68 (26.1) |  |
| 6-10 | 18 (29.5) | 43 (70.5) | 61 (23.4) |  |
| >10 | 52 (45.6) | 62 (54.4) | 114 (43.7) |  |
| **MDA Adverse Effect** |  |  |  |  |
| No | 45 (28.5) | 113 (71.5) | 158 (60.5) | 0.269 |
| Yes | 23 (22.3) | 80 (77.7) | 103 (39.5) |  |

### Community knowledge and self**-**reported preventive behaviors regarding lymphatic filariasis

Overall awareness of LF was high, with 250 of 261 participants (95.8%) reporting that they had heard of the disease. However, only 103 participants (39.5%) demonstrated correct responses across selected indicators of knowledge and preventive behaviors, and MDA participation.

Regarding knowledge of transmission, 123 participants (47.1%) correctly identified mosquito bites as the mode of transmission, while others attributed LF to person-to-person contact, heredity, or spiritual causes. See Fig 2.

**Fig 2.**
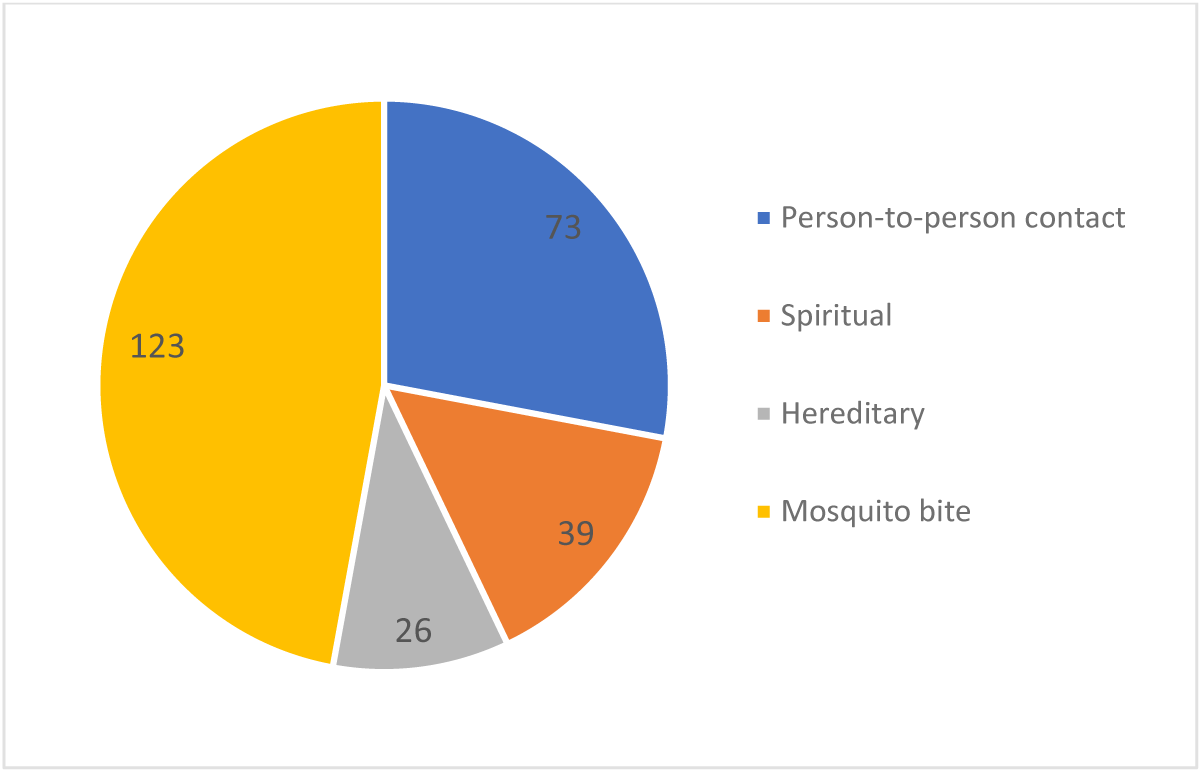
Community knowledge of LF transmission in Birifor, Ghana.

Most participants (185/261; 70.9%) correctly recognized limb or genital swelling as a key clinical manifestation of LF. See Fig 3.

**Fig 3.**
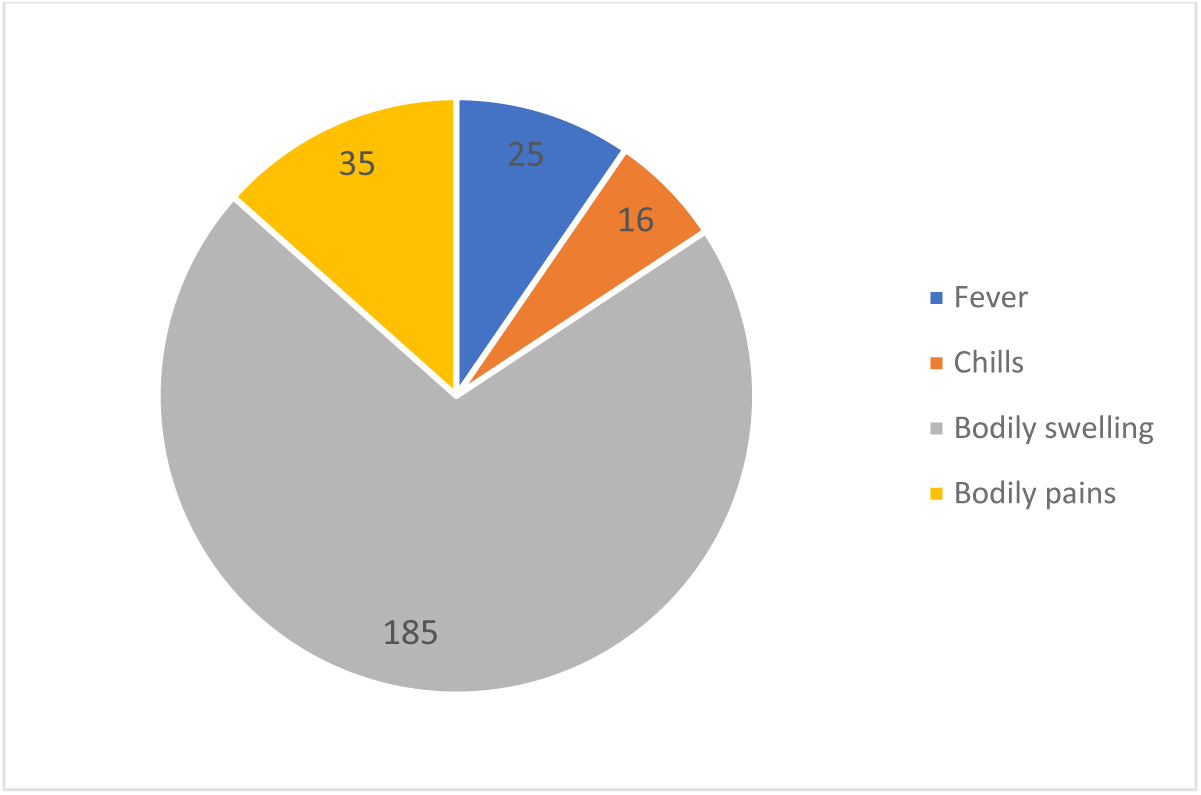
Community knowledge of clinical manifestations of lymphatic filariasis in Birifor, Ghana.

With respect to prevention, 123 participants (47.1%) reported protection against mosquito bites, while 60 (23.0%) mentioned personal hygiene practices as preventive measures. See Fig 4.

**Fig 4.**
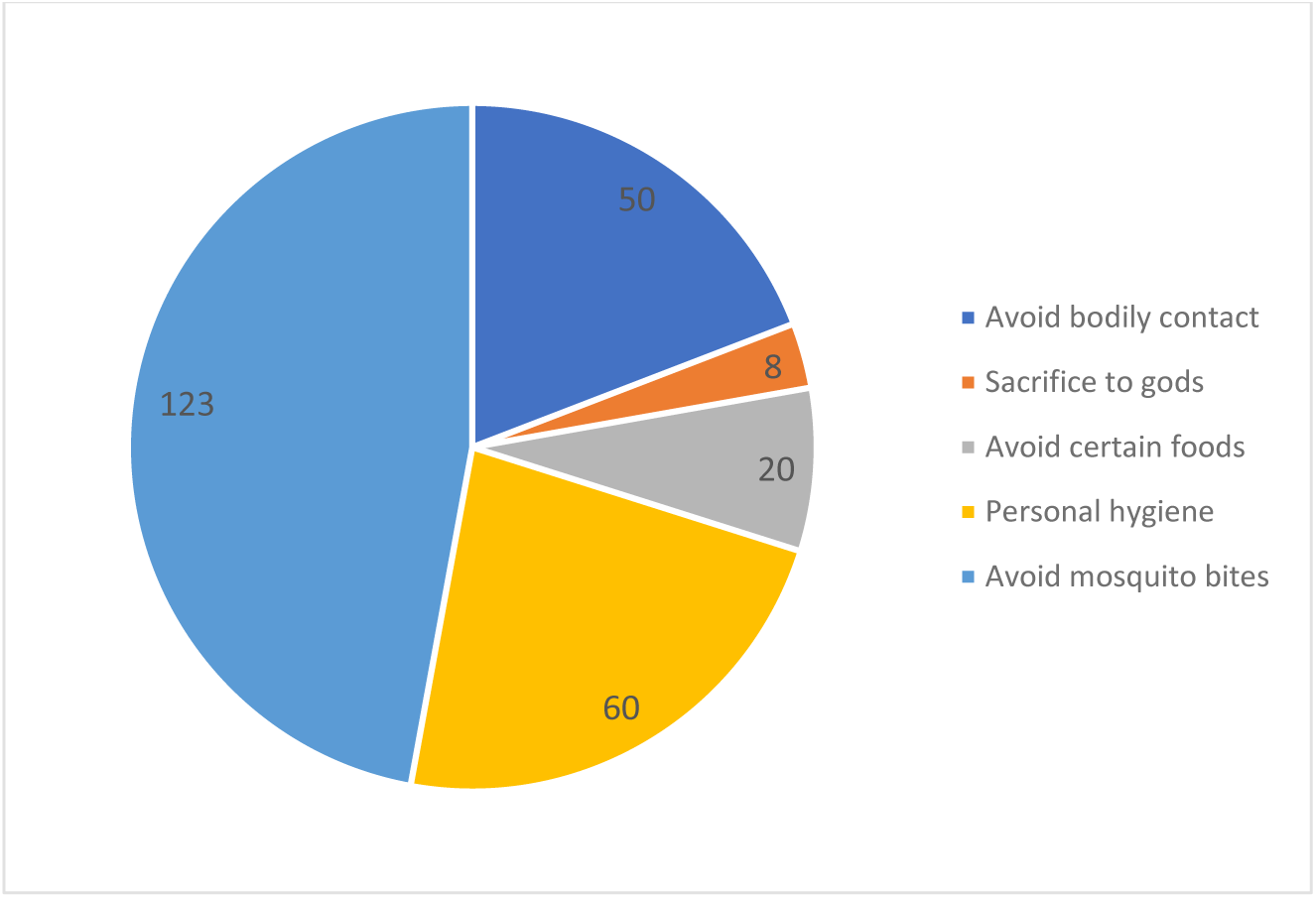
Community knowledge of lymphatic filariasis prevention in Birifor, Ghana.

### Participation in MDA

Awareness of the national MDA program was high, with 250 participants (95.8%) reporting prior knowledge of the intervention. A total of 243 participants (93.1%) reported having ever taken antifilarial medications during MDA campaigns.

Nearly half of the participants (114/261; 43.7%) reported participating in more than 10 rounds of MDA. Most respondents (158/261; 60.5%) reported no adverse effects following treatment. See Fig 5.

**Fig 5.**
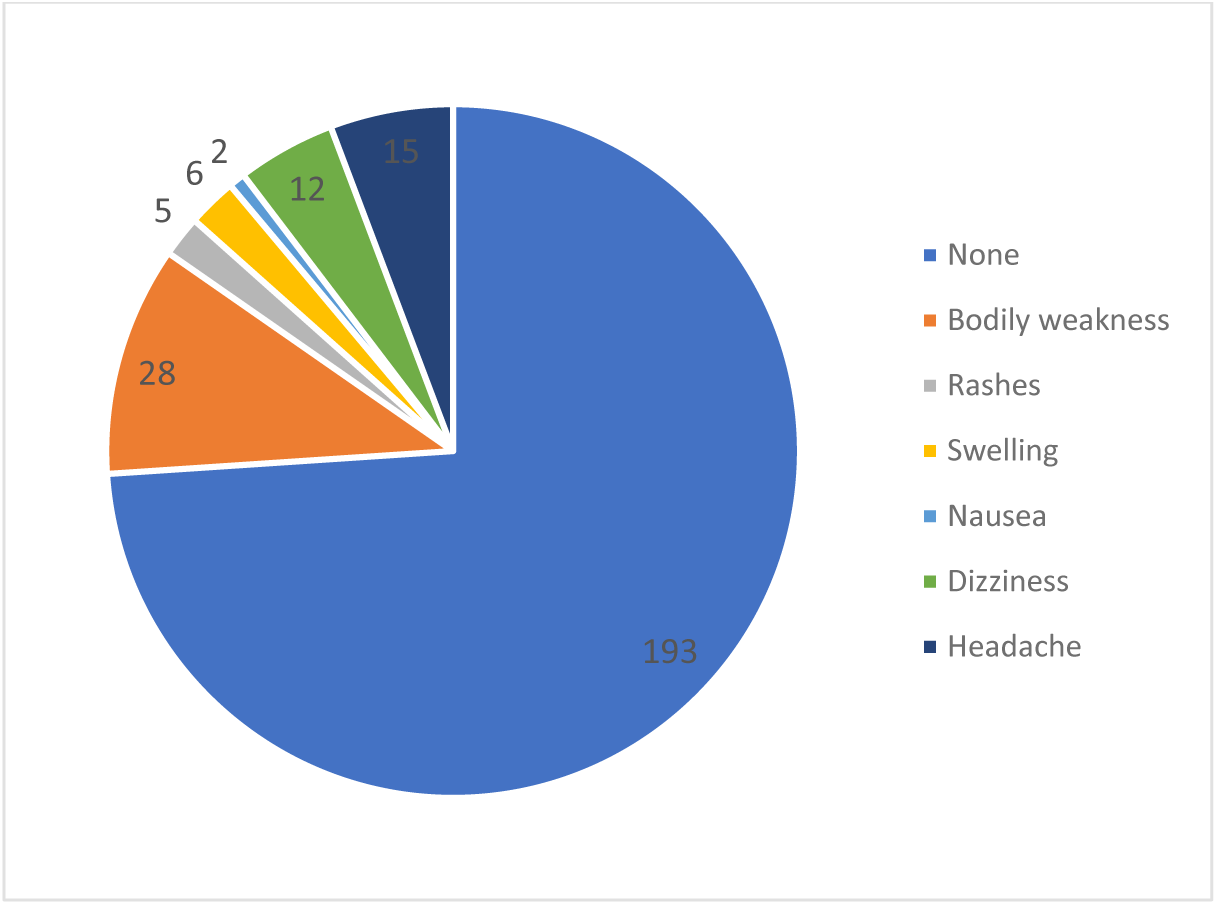
Adverse effects reported after mass drug administration in Birifor, Ghana.

### Prevalence of Microfilariae and Chronic Morbidity

Microscopic examination of night blood smears did not detect microfilariae in any participant (0/261; 0%). Chronic LF-related morbidity was identified in 5 participants, corresponding to a prevalence of 1.9% (5/261). Of these, three cases involved lower limb lymphoedema (all male), while two involved upper limb lymphoedema (one male and one female). All affected individuals were aged above 40 years.

### Impact of Chronic Morbidity on Daily Life

All participants with chronic LF manifestations (5/5; 100%) reported loss of income due to functional limitations associated with the condition. Additionally, 3 individuals (60%) reported that other household members had lost income due to caregiving responsibilities.

### Associations Between Sociodemographic Characteristics and Community Knowledge and Perceptions, and Self-reported Practices

At the bivariate level, several sociodemographic factors were significantly associated with good KAP. This included age greater than 60 years (cOR = 2.57, 95% CI: 1.02–6.47, p = 0.045), being married (cOR = 2.23, 95% CI: 1.31–3.80, p = 0.003), having tertiary education (cOR = 2.83, 95% CI: 1.04–7.72, p = 0.042), and employment in the public sector (cOR = 4.37, 95% CI: 1.43–13.35, p = 0.010).

However, none of these variables remained statistically significant in the multivariable logistic regression model after adjustment for potential confounders (Table 2).

**Table 2.** Factors associated with good knowledge of lymphatic filariasis among community members in Birifor, Ghana.

| Variable | cOR | 95% CI | p-value | aOR | 95% CI | p-value |
| --- | --- | --- | --- | --- | --- | --- |
| **Age** |  |  |  |  |  |  |
| 10-30 | 1 |  |  | 1 |  |  |
| 31-60 | 1.61 | 0.93-2.80 | 0.088 | 1.27 | 0.57-2.83 | 0.552 |
| >60 | 2.57 | 1.02-6.47 | 0.045 | 2.12 | 0.63-7.13 | 0.224 |
| **Marital Status** |  |  |  |  |  |  |
| Never married | 1 |  |  | 1 |  |  |
| Married | 2.23 | 1.31-3.80 | 0.003 | 2.11 | 0.96-4.64 | 0.063 |
| Widowed/Divorced | 1.81 | 0.73-4.49 | 0.202 | 2.68 | 0.77-9.34 | 0.121 |
| **Education Level** |  |  |  |  |  |  |
| No formal education | 1 |  |  | 1 |  |  |
| Primary | 0.62 | 0.33-1.15 | 0.128 | 1.12 | 0.51-2.45 | 0.776 |
| Secondary | 0.98 | 0.51-1.91 | 0.962 | 2.21 | 0.91-5.36 | 0.076 |
| Tertiary | 2.83 | 1.04-7.72 | 0.042 | 4.07 | 0.78-21.21 | 0.096 |
| **Occupation** |  |  |  |  |  |  |
| Student | 1 |  |  | 1 |  |  |
| Farmer | 1.76 | 0.94-3.29 | 0.076 | 1.24 | 0.50-3.08 | 0.647 |
| Trader | 1.4 | 0.64-3.06 | 0.406 | 1.09 | 0.41-2.89 | 0.869 |
| Public sector worker | 4.37 | 1.43-13.35 | 0.01 | 1.45 | 0.27-7.72 | 0.663 |
| Unemployed | 1.79 | 0.37-8.68 | 0.472 | 0.66 | 0.08-5.37 | 0.694 |

## Discussion

This study assessed the epidemiological and community-level status of LF in a rural Ghanaian setting nearly two decades after the implementation of elimination strategies. The absence of detectable microfilariae suggests that transmission is likely very low, although residual infection cannot be excluded, consistent with the long-term impact of MDA programs in endemic areas [12,15]. Similar declines in transmission have been reported in other endemic settings following sustained MDA implementation [20]. However, the persistence of chronic LF-related morbidity (1.9%) underscores that interruption of transmission does not equate to elimination of disease burden.

Despite high awareness of LF (95.8%), only 39.5% of participants demonstrated correct responses across selected indicators of knowledge and preventive behaviors. Misconceptions regarding transmission, particularly beliefs attributing LF to heredity, contagion, or spiritual causes, remain prevalent. These findings are consistent with previous studies documenting persistent misconceptions and cultural interpretations of LF in endemic communities [9,21–23]. While nearly half of the participants correctly identified mosquito bites as the mode of transmission, this partial understanding indicates gaps in health education. Nonetheless, the relatively high awareness of mosquito transmission presents an opportunity to reinforce vector control measures, including the use of insecticide-treated nets, which remain a key preventive strategy [24–26].

The high reported uptake of MDA (93.1%) reflects strong program reach and community engagement. This aligns with evidence from Ghana and other endemic settings demonstrating the effectiveness of repeated MDA rounds in reducing infection prevalence [12,15]. The low incidence of reported adverse effects further supports the safety and acceptability of antifilarial drugs, consistent with findings from other settings where most participants reported minimal or no side effects [12,22,27]. However, even small proportions of individuals who do not participate in MDA may sustain residual transmission [9,11,28–30].

The persistence of chronic morbidity observed in this study is consistent with patterns reported in other endemic regions transitioning toward elimination, where transmission declines but established disease remains [5,6,11,31]. Chronic manifestations such as lymphoedema result from long-term lymphatic damage and may persist even after parasite clearance. The prevalence observed in this study was higher than that reported in some settings [32,33], which may reflect historically higher transmission intensity combined with periods of suboptimal MDA uptake. Importantly, the socioeconomic burden associated with chronic LF was substantial, with all affected individuals reporting loss of income and additional household-level economic strain. Similar findings have been documented elsewhere, where LF-related disability contributes to stigma, reduced productivity, and increased household vulnerability [34–37].

These findings highlight the need to strengthen MMDP as a core component of LF elimination strategies. Evidence shows that simple limb-care practices can reduce acute inflammatory episodes and slow disease progression [38], yet such interventions remain under-prioritized in many endemic settings [31]. Integrating MMDP into routine primary health care systems is essential to ensure sustainability and improve access to care.

From a programmatic perspective, the coexistence of zero detectable microfilaraemia, residual morbidity, and knowledge gaps underscores the need for a dual-track strategy in near-elimination settings. This includes sustaining post-MDA surveillance and addressing the long-term needs of individuals living with chronic LF. Embedding LF indicators into routine health information systems, strengthening community health worker engagement, and implementing targeted behavior change communication strategies are critical to sustaining elimination gains. Similar integrated approaches have been shown to enhance program ownership and long-term impact in neglected tropical disease programs [20].

This study has several strengths. The use of population-based random sampling from an updated census enhances representativeness. The triangulation of parasitological, clinical, and KAP data provides a comprehensive assessment of LF in a near-elimination setting. Additionally, standardized night blood microscopy conducted by trained personnel strengthens the reliability of infection assessment.

However, several limitations should be considered. First, the cross-sectional design limits causal inference and does not capture temporal or seasonal variation in transmission. Second, the use of microscopy alone may have limited sensitivity for detecting low-density infections, particularly in near-elimination settings. As such, the absence of detectable microfilaraemia should be interpreted with caution, as residual infection cannot be definitively excluded without more sensitive diagnostic tools, such as antigen-detection assays or molecular methods [17,39]. Third, the relatively small number of chronic cases limited subgroup analysis. Finally, the selected indicators of knowledge and preventive behaviors used in this study do not constitute a comprehensive KAP assessment, and results should be interpreted accordingly.

The findings of this study are likely generalizable to similar rural, post-MDA settings in northern Ghana and comparable endemic regions, where transmission has declined, but morbidity persists.

Future research should focus on the use of more sensitive diagnostic tools, such as antigen detection assays and molecular methods, to identify residual infection [17,39]. Longitudinal studies are needed to better understand morbidity progression and socioeconomic outcomes. Additionally, implementation research should explore strategies for integrating morbidity management, referral systems, and social support into routine health systems, as well as behaviorally informed interventions to address persistent misconceptions.

These findings highlight the shifting epidemiological profile of lymphatic filariasis in near-elimination settings, where the primary challenges transition from interrupting transmission to addressing residual morbidity, sustaining community engagement, and preventing resurgence.

## Conclusion

This study provides evidence from a rural community in northern Ghana transitioning toward lymphatic filariasis elimination. Although no microfilariae were detected, the persistence of chronic morbidity and substantial gaps in community knowledge highlight ongoing challenges beyond transmission interruption. While awareness of mass drug administration was high, misconceptions regarding transmission and prevention remain common. Chronic lymphatic filariasis continues to impose a measurable socioeconomic burden, underscoring the need for sustained programmatic attention.

Strengthening elimination efforts will require integrated strategies that combine morbidity management and disability prevention, sustained vector control, and targeted social and economic support. Embedding these interventions within routine primary health care systems, alongside continued surveillance and community engagement, will be essential to sustain gains and prevent resurgence in near-elimination settings.

## Data Availability

All data underlying the findings of this study will be made fully available without restriction. The de?identified individual?level dataset and data dictionary will be included as Supporting Information files and will be publicly available upon publication of the article.

## References

1. Azhar S, Naeem M, Qamar W, Iqbal M, Asghar S, Ajaz R, et al. Basic Insights into Lymphatic Filariasis. International Journal of Agriculture and Biosciences. 2023. pp. 73–88. doi:10.47278/book.zoon/2023.53

2. World Health Organization. Integrating neglected tropical diseases into global health and development: fourth WHO report on neglected tropical diseases. Geneva: World Health Organization; 2017. Available: https://www.who.int/publications/i/item/9789241565448

3. Goel TC, Goel A. History. In: Goel TC, Goel A, editors. Lymphatic Filariasis. Singapore: Springer; 2016. pp. 3–5. doi:10.1007/978-981-10-2257-9_1

4. Freitas LT, Khan MA, Uddin A, Halder JB, Singh-Phulgenda S, Raja JD, et al. The lymphatic filariasis treatment study landscape: A systematic review of study characteristics and the case for an individual participant data platform. PLoS Negl Trop Dis. 2024;18: e0011882. doi:10.1371/journal.pntd.0011882

5. World Health Organization. Global programme to eliminate lymphatic filariasis: progress report, 2024. Oct 2025 [cited 3 Apr 2026]. Available: https://www.who.int/publications/i/item/who-wer10040-439-449

6. Medeiros ZM, Vieira AVB, Xavier AT, Bezerra GSN, Lopes M de FC, Bonfim CV, et al. Lymphatic Filariasis: A Systematic Review on Morbidity and Its Repercussions in Countries in the Americas. Int J Environ Res Public Health. 2021;19: 316. doi:10.3390/ijerph19010316

7. Indravudh PP, McGee K, Sibanda EL, Corbett EL, Fielding K, Terris-Prestholt F. Community-led strategies for communicable disease prevention and management in low- and middle- income countries: A mixed-methods systematic review of health, social, and economic impact. PLOS Global Public Health. 2025;5: e0004304. doi:10.1371/journal.pgph.0004304

8. Ahorlu CSK, Koka E, Adu-Amankwah S, Otchere J, de Souza DK. Community perspectives on persistent transmission of lymphatic filariasis in three hotspot districts in Ghana after 15 rounds of mass drug administration: a qualitative assessment. BMC Public Health. 2018;18: 238. doi:10.1186/s12889-018-5157-7

9. Ahorlu CS, Otchere J, Sedzro KM, Pi-Bansa S, Asemanyi-Mensah K, Opare JL, et al. A Comparative Study of Lymphatic Filariasis-Related Perceptions among Treated and Non-Treated Individuals in the Ahanta West Municipality of Ghana. Trop Med Infect Dis. 2022;7: 273. doi:10.3390/tropicalmed7100273

10. Gunaratna IE, Chandrasena NTGA, Vallipuranathan M, Premaratna R, Ediriweera D, de Silva NR. The impact of the National Programme to Eliminate Lymphatic Filariasis on filariasis morbidity in Sri Lanka: Comparison of current status with retrospective data following the elimination of lymphatic filariasis as a public health problem. PLoS Negl Trop Dis. 2024;18: e0012343. doi:10.1371/journal.pntd.0012343

11. Lawford HLS, Tukia ‘Ofa, Takai J, Sheridan S, Ward S, Jian H, et al. Persistent lymphatic filariasis transmission seven years after validation of elimination as a public health problem: a cross-sectional study in Tonga. The Lancet Regional Health – Western Pacific. 2025;57. doi:10.1016/j.lanwpc.2025.101513

12. Angyiereyiri ED, Appawu M, Dadzie S, Boakye D, Otoo S. Impact of Mass Drug Administration (MDA) on the Transmission of Lymphatic Filariasis in Tono Irrigation Area in Navrongo, Ghana. 2015;5: 68.

13. Minetti C, Tettevi EJ, Mechan F, Prada JM, Idun B, Biritwum N-K, et al. Elimination within reach: A cross-sectional study highlighting the factors that contribute to persistent lymphatic filariasis in eight communities in rural Ghana. PLOS Neglected Tropical Diseases. 2019;13: e0006994. doi:10.1371/journal.pntd.0006994

14. Koray MH. Ghana’s path towards eliminating lymphatic filariasis. Trop Med Health. 2024;52: 37. doi:10.1186/s41182-024-00596-2

15. Mensah DA, Debrah LB, Gyamfi PA, Rahamani AA, Opoku VS, Boateng J, et al. Occurrence of Lymphatic Filariasis infection after 15 years of mass drug administration in two hotspot districts in the Upper East Region of Ghana. PLOS Neglected Tropical Diseases. 2022;16: e0010129. doi:10.1371/journal.pntd.0010129

16. Subramanian S, Jambulingam P, Krishnamoorthy K, Sivagnaname N, Sadanandane C, Vasuki V, et al. Molecular xenomonitoring as a post-MDA surveillance tool for global programme to eliminate lymphatic filariasis: Field validation in an evaluation unit in India. PLoS Negl Trop Dis. 2020;14: e0007862. doi:10.1371/journal.pntd.0007862

17. Pastor AF, Silva MR, Dos Santos WJT, Rego T, Brandão E, de-Melo-Neto OP, et al. Recombinant antigens used as diagnostic tools for lymphatic filariasis. Parasit Vectors. 2021;14: 474. doi:10.1186/s13071-021-04980-3

18. Stocks ME, Freeman MC, Addiss DG. The Effect of Hygiene-Based Lymphedema Management in Lymphatic Filariasis-Endemic Areas: A Systematic Review and Meta-analysis. PLoS Negl Trop Dis. 2015;9: e0004171. doi:10.1371/journal.pntd.0004171

19. World Health Organization. Lymphatic filariasis: managing morbidity and preventing disability: an aide-mémoire for national programme managers, 2nd ed. 2021 [cited 3 Apr 2026]. Available: https://www.who.int/publications/i/item/9789240017061

20. Streit T, Lafontant JG. Eliminating lymphatic filariasis: a view from the field. Ann N Y Acad Sci. 2008;1136: 53–63. doi:10.1196/annals.1425.036

21. Kwarteng A, Kenyon KH, Opoku Asiedu S, Garcia R, Kini P, Osei-Poku P, et al. Knowledge and perceptions of lymphatic filariasis patients in selected hotspot endemic communities in southern Ghana. PLOS Glob Public Health. 2023;3: e0002476. doi:10.1371/journal.pgph.0002476

22. Maritim P, Silumbwe A, Zulu JM, Sichone G, Michelo C. Health beliefs and health seeking behavior towards lymphatic filariasis morbidity management and disability prevention services in Luangwa District, Zambia: Community and provider perspectives. PLOS Neglected Tropical Diseases. 2021;15: e0009075. doi:10.1371/journal.pntd.0009075

23. Koly KN, Saba J, Nessa Z, Luba FR, Hossain I, Aktaruzzaman MM, et al. Social and healthcare-seeking experiences of people affected with lymphedema in Bangladesh. PLOS Neglected Tropical Diseases. 2025;19: e0013384. doi:10.1371/journal.pntd.0013384

24. Azzahra M, Shafwah D, Sondakh C, Adriyani R. A Review of Bed Nets Usage and Sewerage Conditions as Risk Factors for Lymphatic Filariasis in Developing Countries. JURNAL KESEHATAN LINGKUNGAN. 2024;16: 89–100. doi:10.20473/jkl.v16i1.2024.89-100

25. Christiana O, Olajumoke M, Oyetunde S. Lymphatic filariasis and associated morbidities in rural communities of Ogun State, Southwestern Nigeria. Travel Med Infect Dis. 2014;12: 95–101. doi:10.1016/j.tmaid.2013.02.006

26. Davis EL, Prada J, Reimer LJ, Hollingsworth TD. Modelling the Impact of Vector Control on Lymphatic Filariasis Programs: Current Approaches and Limitations. Clin Infect Dis. 2021;72: S152–S157. doi:10.1093/cid/ciab191

27. Perera M, Whitehead M, Molyneux D, Weerasooriya M, Gunatilleke G. Neglected Patients with a Neglected Disease? A Qualitative Study of Lymphatic Filariasis. PLOS Neglected Tropical Diseases. 2007;1: e128. doi:10.1371/journal.pntd.0000128

28. Sheel M, Sheridan S, Gass K, Won K, Fuimaono S, Kirk M, et al. Identifying residual transmission of lymphatic filariasis after mass drug administration: Comparing school-based versus community-based surveillance - American Samoa, 2016. PLOS Neglected Tropical Diseases. 2018;12: e0006583. doi:10.1371/journal.pntd.0006583

29. Amanyi-Enegela JA, Kumbur J, Okunade F, Ashikeni D, Ishaya R, Sankar G, et al. Evaluating the effectiveness of mass drug administration on lymphatic filariasis transmission and assessment of post-mass drug administration surveillance in Nigeria’s Federal Capital Territory. Infect Dis Poverty. 2025;14: 63. doi:10.1186/s40249-025-01333-5

30. Srividya A, Dinesh RJ, M M MJ, Kishanthini M, Dogra V, Tripathi B, et al. Cross-sectional epidemiological assessment of lymphatic filariasis situation in areas under post-mass drug administration surveillance and the associated risk of transmission in the context of migrants in India: a study protocol. BMJ Open. 2026;16: e116111. doi:10.1136/bmjopen-2025-116111

31. Hafiz I. Investigation of lymphatic filariasis distribution, morbidity management and disability prevention in Bangladesh. University of Liverpool. 2018.

32. Adekunle NO, Asimiea AO. Prevalence of Lymphatic Filariasis and Associated Clinical Morbidities Among Adolescents in Three Rural Communities in Ondo State, Southwest Nigeria. Journal of Tropical Medicine and Health. 2018. doi:Adekunle O. Prevalence of Lymphatic Filariasis and Associated Clinical Morbidities Among Adolescents in Three Rural Communities in Ondo State, Southwest Nigeria. Journal of Tropical Medicine and Health. doi:10.29011/JTMH-120.000120

33. Jian H, Lawford H, McLure A, Lau C, Craig A. Global Lymphatic Filariasis Post-Validation Surveillance Activities in 2025: A Scoping Review. Tropical Medicine and Infectious Disease. 2026;11: 28. doi:10.3390/tropicalmed11010028

34. Gyapong JO, Gyapong M, Evans DB, Aikins MK, Adjei S. The economic burden of lymphatic filariasis in northern Ghana. Ann Trop Med Parasitol. 1996;90: 39–48. doi:10.1080/00034983.1996.11813024

35. Krishna Kumari A, Harichandrakumar KT, Das LK, Krishnamoorthy K. Physical and psychosocial burden due to lymphatic filariasis as perceived by patients and medical experts. Trop Med Int Health. 2005;10: 567–573. doi:10.1111/j.1365-3156.2005.01426.x

36. Akinreni T, Akinreni M, Adebowale KA, Azubuike PC, Frank IU, Ogundipe O, et al. Lived experiences, stigma and impacts of lymphatic filariasis: a scoping review. Int Health. 2026; ihag027. doi:10.1093/inthealth/ihag027

37. Asiedu SO, Kwarteng A, Amewu EKA, Kini P, Aglomasa BC, Forkuor JB. Financial burden impact quality of life among lymphatic Filariasis patients. BMC Public Health. 2021;21: 174. doi:10.1186/s12889-021-10170-8

38. Wijesinghe RS, Wickremasinghe AR, Ekanayake S, Perera MSA. Efficacy of a limb-care regime in preventing acute adenolymphangitis in patients with lymphoedema caused by bancroftian filariasis, in Colombo, Sri Lanka. Ann Trop Med Parasitol. 2007;101: 487–497. doi:10.1179/136485907X193806

39. Rocha A, Braga C, Belém M, Carrera A, Aguiar-Santos A, Oliveira P, et al. Comparison of tests for the detection of circulating filarial antigen (Og4C3-ELISA and AD12-ICT) and ultrasound in diagnosis of lymphatic filariasis in individuals with microfilariae. Mem Inst Oswaldo Cruz. 2009;104: 621–625. doi:10.1590/s0074-02762009000400015

